# Investigating the Impact of Prediabetes on the Aging Process through Longitudinal Analysis of Blood-Based Biomarkers: A Systematic Review and Meta-Analysis Protocol

**DOI:** 10.1101/2023.07.31.23293409

**Authors:** Nonkululeko Avril Mbatha, Nomusa Christina Mzimela, Aganze Gloire-Aimé Mushebenge, Andile Khathi

## Abstract

**Background:** Prediabetes is a disorder that affects the metabolic function of the body, and it can lead to heart disease, stroke, and type 2 diabetes (T2D). Previous studies have reported a correlation between T2D and exacerbated senescence, however, none have reported a link between prediabetes and senescence. Hence, this systematic review protocol and meta-analysis will be the first, to the best of our knowledge, to provide detailed guidance on all steps taken in the synthesis and meta-analysis of data reporting the correlation of prediabetes with senescence by identifying changes to biological aging indices.

**Methods and analysis:** The PRISMA 2015 reporting protocol preparation standards were followed in the creation of this protocol. The search for pertinent studies will be undertaken by the framework for Arksey and O’Malley reviews and the Preferred Reporting Items for Systematic Reviews and Meta-Analyses (PRISMA-ScR). Clinical studies published in English from January 2003 to March 2023 with observational methods will be used, including cross-sectional, comparative cross-sectional, case-control, and cohort study designs with normal/non-diabetic, prediabetic, and type 2 diabetic cases reporting on senescence and changes in blood-based biomarker levels in a multi-ethnic population aged 25-45 years. An extensive search of relevant studies will be conducted in the following databases: PubMed, Google Scholar, Cochrane Library, ScienceDirect, Web of Science, Scopus, WHO Global Health Library, and African Journals Online. In addition, all the results will be examined for eligibility by two reviewers (NAM, NCM). Any differences between the two authors will be settled by a third reviewer (GAM), to ensure the overall quality of the findings. To find additional relevant studies, authors will also look through reference lists, grey literature sources, and peer-reviewed journals. The risk of bias will be examined using the Downs and Black checklist. To assess statistical heterogeneity of the studies, a standard chi-square test will be used with a significance level of P 0.10 indicating that there is no true heterogeneity between the studies. For the meta-analysis and to analyse the sensitivity of the data, Review Manager (RevMan) software (version 5.4) will be used to populate forest plots that will display the effect estimates and confidence intervals from each study. The strength of the evidence will be evaluated using the Grading of Recommendations Assessment, Development, and Evaluation (GRADE) system.

**Results and Conclusion:** This protocol aims to provide guidance on how to investigate articles that reported on the correlation of prediabetes with exacerbated senescence by identifying common pathways utilized and changes induced to biomarkers of biological aging. The results from this protocol will highlight research gaps in the impact of prediabetes on aging and provide suggestions for future research. In addition, it will provide evidence-based information to give guidance to policymakers on treatment decisions to improve patient outcomes.

**Ethics and Dissemination:** No ethical approval is required as the data under consideration have already been published and no additional data will be requested from participants. The results of this review will be disseminated through a peer-reviewed publication and presented at pertinent conferences.

**Registration Details:** The International Prospective Registry of Systematic Reviews (PROSPERO) has been used to register this protocol, registration number (CRD42023407084) dated 05/04/2023.

## 1. Background

Prediabetes a metabolic condition that is characterized by moderate glycaemic dysregulation and is a frontline risk factor for multiple metabolic complications such as type 2 diabetes (T2D), heart diseases and stroke if left untreated (1). The condition is characterized by blood glucose levels that are above the upper threshold of normal but below the threshold of T2D (1). Individuals with glycated haemoglobin (HbA1c) of 5.7–6.4% or impaired glucose tolerance (IGT) of 7.8-11.0 mmol/L (140-200 mg/dL) after 2 hours post consumption of 75g of oral glucose load or impaired fasting glucose (IFG) of 6.1-6.9 mmol/L (110 to 125 mg/dL) are verified to be prediabetic either by World Health Organisation (WHO) and the American Diabetes Association (ADA) (2,3). Hence, the WHO and ADA criteria are typically utilized when calculating the prevalence of prediabetes. Globally, the prevalence of prediabetes was estimated to be 7.5% (374 million) in 2019, and it is anticipated to increase to 8.0% (454 million) by 2030 then approximately reach 8.6% (548 million) by 2045 (4). According to Stats SA, the prevalence rate of diabetes mellitus among the South African population is 11.3% and is the leading cause of death among women (5). Shockingly, over 50% of the South African population still remains undiagnosed with this condition. In 2019, South Africa was designated as the “unhealthiest country” by the Indigo Wellness Index, primarily due to unfavourable scores obtained in the assessments of obesity and other indicators related to diabetes risk (6). Intriguingly, the prevalence of prediabetes in South Africa was reported to have almost doubled from 35% in 2019 to 65% in 2020 and it is currently at 67% from 2022 (7). Furthermore, there is evidence indicating that prediabetes exhibits a greater prevalence among individuals within the middle-aged group (25-45 years old) (8-10). Moreover, there is a notable trend of prediabetes progressing to T2D with advancing age, as the older-aged group (45-60 years old) displays a higher prevalence of T2D (8, 11-13). Furthermore, studies conducted in mixed ethnic populations reported that prediabetes prevalence is disproportionally higher in certain ethnic groups due to genetic predisposition (14-17). Hence, this review selected a multi-racial, middle-aged group (25-45 years old) to investigate the intersectional impact of prediabetes in biomarkers of aging and a comparison to non-diabetic individuals of the same criteria will be conducted.

Previous research has reported a significant correlation between T2D and the facilitation of biological aging (BA), also known as senescence. This association is supported by findings of increased telomere shortening and mitochondrial DNA depletion, accelerated aging of human collagen, and the induction of senescence leading to accelerated vascular aging in type 2 diabetic individuals (18-22). Furthermore, BA is hallmarked by cellular senescence, which arises from the gradual decline in the functional systems of the human body (23-25). This phenomenon is also observed as a pathophysiological feature in T2D, emphasizing the intricate interplay between T2D and the ongoing process of progressive aging. This also emphasizes the complex relationship between metabolic disorders and BA.

Theoretically, it is recognized that prediabetes may play a role in promoting the aging process through mechanisms involving oxidative stress, inflammation, and cellular damage. However, there remains a significant research gap in comprehending how prediabetes precisely facilitates the aging process and induces alterations in aging biomarkers. This knowledge gap arises due to the complexity of the process, which encompasses multiple interconnected biological pathways.

Currently, there is a dearth of studies establishing a correlation between prediabetes and the facilitation of BA. Moreover, there is a lack of evidence from intersectional studies that specifically examine blood-based biomarkers of aging in individuals with prediabetes and compare these findings with those of non-diabetic individuals. Therefore, it is strongly recommended to conduct further research studies on this topic in order to obtain a more comprehensive understanding of the impact of prediabetes on senescence and to precisely identify the extent of its involvement in the aging process. The discovery of these underlying processes could potentially lead to the development of novel personalized treatment strategies aimed at delaying or even halting the aging process in individuals with prediabetes, thereby potentially enhancing the effectiveness of interventions and improving health outcomes. Furthermore, these findings have the potential to facilitate early intervention in prediabetes and mitigate its progression to aging-related diabetic complications, including cardiovascular complications.

The calculation of BA with precise accuracy remains a challenge, as no single biomarker can provide a comprehensive assessment. However, previous studies have reported elevated levels of various aging markers in the senescent state of individuals with T2D when compared to non-diabetic groups. These markers include Cyclin-dependent kinase inhibitor 2A (CDKN2A), also known as p16INK4a, Senescence-associated β-galactosidase (SA-gal), reactive oxygen-containing species (ROS), telomere shortening, Interleukin 6 (IL-6), and Tumour Necrosis Factor Alpha (TNF-α) (18, 26-31). The observed elevation of these markers in individuals with type 2 diabetes indicates a plausible link between diabetes-induced senescence and the dysregulated expression of these markers, thereby contributing to the progression of the aging process. These findings underscore the intricate and multifaceted nature of the interplay between diabetes and the senescence-associated aging process. Consequently, this gave rise to the interest to conduct a systematic review that assesses the mechanism utilized by prediabetes to facilitate BA while also identifying the involvement of all blood-based biomarkers of aging. The aim is to achieve a thorough synthesis, based on studies that have already been gathered and reported on the changes induced by aging biomarkers due to prediabetes and identify its mechanism of involvement in the aging process. Furthermore, this enabled this study to generate the following objectives.

### Objectives

1. To examine the role of prediabetes on the biological aging process by identifying alterations induced to blood-based biomarkers of aging in a multi-ethnic population aged 25-45 years..
2. To identify the specific mechanisms by which prediabetes facilitates the biological aging process and determine its level of involvement.
3. To map the intersectional differences of biological aging, by conducting a comparative analysis in individuals experiencing healthy aging compared to that undergoing prediabetes influenced aging.

## 2. Methods and Design

The guidelines from preferred reporting items for systemic reviews and meta-analysis (PRISMA), 2015 were adhered to during the preparation of this protocol (An additional file of the PRISMA checklist has been included).

### 2.1. Systematic Review Registration

The International Prospective Registry of Systematic Reviews (PROSPERO) has been used to register this protocol, (PROSPERO registration number “CRD42023407084” dated 05/04/2023).

### 2.2. The Study’s Eligibility Criteria

Studies reporting community-based clinical cross-sectional studies with a minimum of 100 participants will be considered, to ensure that this review is comprehensive and unbiased. The following inclusion and exclusion criteria will be applied:

#### Inclusion criteria

Studies/articles published from 2003 to 2023, that report on prediabetes, aging, and blood-based biomarkers of aging in a multi-ethnic population will be considered.

#### Exclusion criteria

*S*tudies that were published before 2003 with a sample size of less than 100 subjects that do not report on human subjects, prediabetes, aging, and biomarkers of aging will be excluded. Non-primary research studies like reviews, editorials, and studies that have incomplete or unclear data reported will also be excluded.

### 2.3. Criteria for Prediabetes Diagnosis

Individuals should meet the following diagnosis criteria, high glycated haemoglobin (HbA1c) of 5.7–6.4% or impaired glucose tolerance (IGT) of 7.8-11.0 mmol/L (140-200 mg/dL) after 2 hours post consumption of 75g of oral glucose load or impaired fasting glucose (IFG) of 6.1-6.9 mmol/L (110 to 125 mg/dL) will be verified to be prediabetic as per the WHO and the ADA prediabetes diagnosis criteria (2,3). Also, prediabetes diagnosis with HbA1c will be accepted for any value between 5.7% and 6.5% (3).

### 2.4. Study Design

To ensure optimal reliability of the desired findings and enhance statistical significance, any clinical studies that reported on prediabetes and aging from 2003 to 2013, consisting of a minimum of 100 subjects from all genders that are multi-racial will be the focus for the source of information. To maximize the potential of acquiring fundamental concepts of this review, the PICO framework (32) as tabulated in Table 1 will be implemented as follows.

**Table 1:** PICO Framework.

| **Framework** | **Evidence-based practice** |
| --- | --- |
| **P: Population** | Prediabetic subjects that are multi-ethnic, from both genders, and aged 25-45 years old. |
| **I: Intervention** | None as this is an observational study. |
| **C: Comparison** | Individuals that are healthy and non-prediabetic will be used as the control group. |
| **O: Outcome** | Role of Prediabetes in Aging and Changes Induced to blood-based biomarkers of Aging. |

#### 2.4.1. Research Questions

The following research questions were developed to improve our understanding of the impact of prediabetes on aging,

1. What is the impact of prediabetes on aging and what changes does it induce to blood-based biomarkers of aging?
2. What is the involvement of prediabetes on aging and what mechanism does it use to facilitate aging?
3. What intersectional inequalities can be identified in blood-based biomarkers of healthy aging when compared to prediabetic-induced aging?

## 3. Outcomes

The expected outcomes of this systematic review are as follows.

### Primary outcomes

- The impact of prediabetes on the aging process and alterations induced to biomarkers of aging.
- The mechanism utilized by prediabetes to facilitate aging and its involvement in the aging process.
- Changes in biomarkers of senescence indices associated with prediabetes during aging.

### Secondary outcome

Intersectional differences in biomarkers of aging between healthy aging versus prediabetes-associated aging.

#### 3.1. Search strategy

With a librarian’s aid, two independent reviewers (NAM, NCM) will conduct a comprehensive search of databases to find all related articles published on, prediabetes, the human aging process, and blood-based biomarkers of aging that were published from 2003 to 2023, regardless of the language of publication. The databases that will be screened will include MEDLINE through PubMed, Google Scholar, WHO Global Health Library, CHOCHRANE, Web of Science, EMBASE, and African Journals Online. They will use some of the following medical subject headings in our search strategy: “Prediabetes and biological aging of the human body,” “blood-based biomarkers of aging,” “mechanisms utilized by prediabetes to facilitate aging” “intersectional differences in aging induced by prediabetes”. EndNote referencing manager will be used to remove duplicates. In addition, a hand search will be conducted to identify other eligible studies that are not indexed in these databases, using the bibliography of included studies and relevant literature reviews.

#### 3.2. Types of eligible studies

Clinical studies published in English from 2003 to 2023 with observational methods will be used, including cross-sectional, comparative cross-sectional, case-control, and cohort study designs with normal/non-diabetic, prediabetic, and type 2 diabetic cases reporting on senescence and changes in blood-based biomarker levels in a multi-ethnic population aged 25-45 years. The study must have a minimum of 100 participants for it to pass eligibility and subjects must be over 18 years of age and be clinically diagnosed with prediabetes according to the prediabetes diagnosis criteria defined by the ADA and the WHO (2,3). Any study lacking a clear diagnosis criteria description will be excluded if, after contacting authors twice, the information is not provided. There will be no language restrictions and the most recent version of the study will be selected if there are multiple articles reporting the same results.

#### 3.3. Patient and Public Involvement

There will be no patient involvement.

#### 3.4. Data Management

### 3.4.1. Data extraction

Using a predesigned Excel form, the reviewer (NAM) will extract the applicable data. To ensure the quality of extracted data, another reviewer (NCM) will independently check all data. A third reviewer (GAM) will resolve any disagreements between the two authors to ensure a thorough and unbiased evaluation of the relevant studies and to enhance the overall quality of the findings. The data to be extracted will include the population being sampled, prediabetes, and biological aging blood-based biomarkers reported which will be stratified by age, sex, and ethnicity. Data on family history parameters such as weight, hypertension, and diabetes will be pulled to appraise the most conventional risk factors associated with prediabetes. The categories (period, design, sampling approach, and a distinction between whether participants are prediabetic or non-prediabetic will also be taken into consideration along with the methodology of the study described. Lastly, once all the inclusion criteria elements have been satisfied, extraction of the data will be executed. Studies with insufficient data to calculate the primary outcome will face exclusion if the requested data is not provided after contacting the corresponding author twice.

### 3.4.2. Data Condensation

All the pertinent studies that will be included, will be grouped into two main groups according to the diagnosis of participants whether they are prediabetic or non-prediabetic. Then four subgroups will be done according to ethnicity (Black, Indian, White, Colored). Lastly, the data will further be divided into5 sub-groups according to participants’ age as follows: (group 1, 25-29 years old; group 2, 30-34 years old; group 3, 35-39 years; group 4, 40-45 years old).

#### 3.5. Risk of Bias

The risk of bias will be examined using the Downs and Black checklist (33). To assess the quality of a study, the following score rating will be applied; (26-28 points are excellent, 20-25 points are good, 14-19 points are fair, 8-13 points are poor, and 0-7 points are very poor). Quality assessment will be undertaken by two reviewers independently (NAM, NCM). Disagreements will be resolved by discussion, and a third reviewer (GAM) will arbitrate judgments on the overall risk of bias. The risk of bias will be categorized as either low, moderate, or high risk according to the following domains: reporting (10-12 items), external validity (3-5 items), internal validity (11-14 items) and bias (6-8 items).

#### 3.6. Data synthesis and analysis

The RevMan software version 5.4 will be used for the meta-analysis since it enables the pooling of effect estimates from many studies and the creation of a forest plot to show the outcomes graphically (34,35). Forest plots will offer a graphical depiction of the meta-analysis’s findings of the impact induced by prediabetes on aging. This will include the effect sizes of the individual studies, their respective confidence intervals, and the estimated total pooled effect size. Each research finding will be represented as a square or diamond on the plot, along with the effect size and its confidence interval. Each square or diamond’s size reflects the importance of the study to the meta-analysis. The study-specific estimates with a 95% confidence interval will be pooled to generate an overall summary of the impact of prediabetes on blood-based biomarkers of aging figures across the selected studies.

#### 3.7. Analysis of Sensitivity

To assess statistical heterogeneity of the studies, a standard chi-square test will be used with a significance level of P 0.10 indicating that there is no true heterogeneity between the studies (36). For the meta-analysis and to analyze the sensitivity of the data, RevMan version 5.4 will be used to populate forest plots that will display the effect estimates and confidence intervals from each study.

#### 3.8. Confidence in cumulative evidence

The Grading of Recommendations Assessment, Development, and Evaluation (GRADE) method will assess the strength of evidence (37,38). The GRADE method will provide a score of the quality of the studies and the strength of the evidence depending on methodological flaws within the included studies, consistency of results across diverse studies, precision estimates, and publication bias. This decision will be made separately by two reviewers (N.A.M. and A.K.).

## 4. Discussion and conclusion

This systematic review protocol was created to offer a thorough and rigorous method for a systematic review and meta-analysis on investigating how prediabetes affects aging and the alterations induced to aging-related blood-based biomarkers. The aim of synthesizing existing research is to identify the specific biological mechanisms and blood-based biomarkers involved in prediabetes-induced aging. The outcomes from this systematic review analysis uncover knowledge gaps and offer new insights into the intricate relationships between prediabetes and aging. Furthermore, the findings of this systematic review protocol also highlight the need for further research to be conducted on the relationship between prediabetes and aging while highlighting significant implications for the development of targeted interventions and personalized strategies to improve health outcomes for individuals with prediabetes. In addition, the results from this systematic review will be disseminated through presentations at conferences, peer-reviewed journals, and other pertinent venues as well as the public.

## Author’s contributions

N.A.M., AGAM and A.K. were responsible for brainstorming, designing the study, and drafting the protocol. N.A.M., G.A.M., N.C.M., and A.K. were responsible for reviewing study eligibility, checking bias, and finalizing the published version of the manuscript. All authors have read and agreed to the published version of the manuscript.

**Correspondence**: Nonkululeko Avril Mbatha, **Twitter** @avrilmbatha

## List of Abbreviations

T2D: Type II Diabetes
GRADE: Grading of Recommendations Assessment, Development, and Evaluation
WHO: World Health Organization
ADA: American Diabetes Association
PICO: Population, Intervention, Comparison, and Outcomes
BA: Biologic aging, PRISMA preferred reporting items for systemic reviews and meta-analysis
RevMan: Review Manager
BA: Biological aging.

## Institutional Review Board Statement

There will be no need for signed informed permission and ethics approval as the systematic review and meta-analysis will analyze data that are published.

## Informed Consent Statement

Not Applicable.

## Data Availability statement

Since it is a protocol for a systematic review, there are no more data accessible than the file that is provided.

## Acknowledgments

The authors are appreciative of the University of KwaZulu Natal, College of Health Science.

